# The burden of pertussis disease and vaccination coverage in Australian adults attending primary health care

**DOI:** 10.1101/2025.08.09.25333139

**Authors:** Aye M. Moa, Juan C. Vargas-Zambrano, Hubert Maruszak, Valentina Costantino, C. Raina MacIntyre

## Abstract

**Background:** The reported incidence of pertussis, a vaccine-preventable disease, has been increasing in recent years. This study aimed to estimate the burden of pertussis and vaccination rate in Australian adults in primary care.

**Methods:** Deidentified data for subjects ≥18 years were extracted from Medical Director (MD) primary care software from 2008-2019. We estimated the cumulative incidence of diagnosed pertussis in adults by age and risk groups, and vaccine coverage in cases and a control group (not diagnosed with pertussis or a coughing illness). We also examined the incidence of undiagnosed coughing illness in the study population.

**Results:** Of the 764,864 subjects included in the study, 1,788 (0.2%) were diagnosed with pertussis between 2008 and 2019, corresponding to an average annual diagnosis rate of 76.9 per 100,000 population. About 31,110 (4.1%) of adults had an undiagnosed coughing illness. The highest rate was observed in 2011 and higher in females (63.3%), and the diagnosis rate was stable across all age groups. Underlying chronic conditions were more prevalent among pertussis cases than controls (58.7% vs 18.8%), with asthma or chronic obstructive pulmonary disease (COPD) being the most common. Overall, 14% of cases received a pertussis vaccination during the study period. Diagnostic testing for pertussis was performed in 34.1% of pertussis cases. Estimated conservative costs per pertussis patient ranged from AUD $473 to $909, with higher costs observed in individuals with complications.

**Conclusion:** In the outpatient setting, there was a notable burden of pertussis among adults under 65 years of age, particularly those with underlying medical conditions, such as asthma and COPD which appear to be significant risk factors. Due to low rate of pertussis testing among all coughing illness, a proportion of non-specific coughing illness may be undiagnosed pertussis. The observed low vaccination rates highlight a need for increased awareness, improved diagnostic efforts and prevention strategies in primary care.

## Background

Pertussis is a vaccine preventable disease that affects all ages but is the most severe in infants. However, older adults and people with chronic illnesses such as asthma or COPD have higher morbidity and mortality compared to healthy adults.^1,2^

Generally, outbreaks of pertussis occur every 2-5 years, and prior to COVID-19, it was the second highest infectious disease notified to National Notifiable Diseases Surveillance System (NNDSS) in Australia.^3^ Australia has had among the highest reported incidence of pertussis globally in some years,^4^ with large outbreaks in 2011 and 2024, with a notification rate of 94.8 and 215 per 100,000 population, respectively. Between 2013 and 2018, the average all-age pertussis notification rate per year in Australia was 63.6 per 100,000 population, with a shift in burden to adults.^3^ During 2020-2022, corresponding to COVID-19 mitigation measures, pertussis was much lower than pre-pandemic years, however, incidence has increased since 2023, with the largest epidemic ever reported in 2024.^5^ Probable and confirmed pertussis cases are notifiable in Australia, but testing and awareness of pertussis is low in adults, ^6^ with underdiagnosed and under-reporting by GPs.^7^ In contrast to children, the clinical presentation in adults can be atypical, often presenting with a non-specific or atypical cough, and general practitioners (GPs) may not test for pertussis.^8-10^ Severe disease and deaths may occur in older adults, and outbreaks can occur in aged care facilities.^11^ Identification of pertussis depends on health seeking by the patient, GP’s awareness of the disease and diagnostic testing.^12^ Further, there are few GP consultations for pertussis vaccination and limited knowledge about the severity of the disease in adults.^12^ Further, testing might not be conducted appropriately with either PCR ordered too late or serology ordered too early, which may contribute to misdiagnosis. Opportunistic pertussis vaccination is recommended in the Australian Immunisation Handbook when a tetanus booster is required, but about one third of Australians receive dT instead of dTpa.^13^

The pertussis vaccine is recommended but not funded for adults ≥65 years when their last dose was more than 10 years ago, or for any adult requesting the vaccine. ^14^ It is also recommended for at-risk populations such as healthcare workers, early childhood educators, and those providing care to children, including new parents.^14^ The pertussis vaccine is available only in combination form with tetanus and diphtheria, for adolescents and adults in Australia.^14^ Pertussis vaccination in adults remains suboptimal nationally.^15^ Data from the Australian Immunization Registry in 2023 showed that 21% of adults aged 50 years or older were up-to-date for pertussis vaccination compared to tetanus and diphtheria vaccination which was about 30%.^13^ Australian pertussis vaccination rates are lower than in the United States, where overall Tdap vaccine coverage in adults aged >19 years was 28.6%.^16^ There is limited information on pertussis disease burden in primary care in Australian adults. This study aimed to determine the diagnosis of pertussis disease, associated complications and the vaccination rate in Australian adults ≥18 years of age attending primary health care.

## Methods

We conducted a retrospective descriptive study in patients ≥18 years attending the practices of consenting clinicians for a 12-year index period (2008-2019).

### Data source

Study data were extracted electronically using MedicalDirector (MD), a national clinical practice management software system used by just under half of all Australian GPs. As of December 2024, the database consisted of approximately 1,300 users; primarily primary care practitioners (90-95%) and a smaller proportion of specialists. Participating clinicians had consented to participate and provide de-identified, and aggregated patient data for research purposes. Geographically, the majority of data came from New South Wales and Victoria (∼30-35%), followed by Queensland (<20%) and other states comprised <10%. All analyses followed MD’s data governance model, which is underpinned by five tenets: integrity, safety, control, utility and transparency. Data output was subject to minimum aggregation requirements; whereby no single cell of any output data was provided where the underlying data had 10 or fewer patients or patients from 3 or fewer medical practices due to privacy and confidentiality issues.

### Study population

The sample frame included all adults aged 18 years and over, who had attended one of the primary care sites which use MD and where the clinician had consented. As patients are deidentified, if they attend more than one clinic, their records in each clinic appear as separate records. To be eligible for the study, we identified a targeted sample of “active patients”, defined as those who regularly attend a single clinic, and analysis was restricted to active patients only. We used the Royal Australian College of General Practitioners definition of an active patient - an individual who has attended the same practice a minimum of 3 times in the previous 2 years.^17^ The population at risk was considered as those active patients for each calendar years as described below. Deidentified, aggregated data was extracted electronically from participating primary care clinics over the study period (01 January 2008-31 December 2019), aimed to provide data broadly representative of the national primary care patient population.

### Study sample

Data were collected on patients with diagnosed pertussis and with unspecified coughing illness (not diagnosed with pertussis), as well as a control group of patients attending clinics, who met the selection criteria to compare with pertussis cases for vaccination, clinical and sociodemographic characteristics. Details of selection criteria for each group are presented below. The study period was from 2008 to 2019.

### Diagnosed pertussis cases

Pertussis cases were defined as patients who had a documented diagnosis of pertussis in their medical record in the study period, with or without laboratory confirmation of pertussis. If pertussis diagnosis was noted more than once for the same patient, then we only included the first diagnosis recorded in their patient file.

### Undiagnosed coughing illness

A secondary outcome was the incidence of coughing illness to determine the potential burden of coughing illness that may be related to undiagnosed pertussis in the study cohort attending primary care. Cases of coughing illness were defined as patients who did not have diagnosed pertussis and have had a coughing condition with no other diagnosis (as shown in the Supplementary, Table S2) at any point during the study. Patients who have had coughing illness with a known, diagnosed condition were excluded (n= 5016 patients). These include gastro-oesophageal reflux disease (GERD), allergic rhinitis, chronic rhinitis, chronic sinusitis, interstitial lung disease, lung cancer, chronic cough (with asthma), chronic cough (with COPD) and chronic cough (in patients who had been prescribed with angiotensin-converting enzyme (ACE) inhibitors), and coughing condition due to any other respiratory infections, i.e. influenza or RSV.

#### Controls

Unmatched controls were selected by identifying active patients in the study period who did not have pertussis nor coughing illness during the study period.

### Study outcome and data analysis

The primary outcomes of interest were the cumulative incidence of pertussis or coughing illness in the eligible population of adults aged ≥18 years during the study period (2008-2019). Study data were analysed using descriptive epidemiology to compare three groups: patients diagnosed with pertussis, those with an unspecified coughing illness, and controls with neither condition. The overall study population and population subgroups were calculated using a denominator of all ‘active patients’ (overall and within each subgroup). This allowed the calculation of cumulative incidence for the overall population and each subgroup. We stratified age by three groups, 18-44 years, 45-64 years, ≥65 years. We also analysed data by at-risk conditions such as asthma, chronic obstructive pulmonary disease (COPD), cardiovascular disease (CVD), diabetes and obesity. For analysis by states, data regarding state and territory there were some limitations. There was missing data or limited information on post codes and data were not available for Australian Capital Territory and Northern Territory due to values <10 counts in a cell.

Pertussis vaccination rate was estimated across all three groups among individuals who had received pertussis vaccine as adults at any point in the study period. The timing of vaccination was also estimated as being in the last 5 years (2015-2019) or earlier (2008-2014). In addition, patient records were reviewed to determine the timing of pertussis vaccination in relation to diagnosis – specifically, whether the vaccine was administered before or after the diagnosis. Data on other adult vaccines such as influenza, pneumococcal, and herpes zoster, administered during the study period were also collected for comparison.

We determined the number of patients who had laboratory testing for pertussis using polymerase chain reaction (PCR) or serology and the number of prescriptions issued for patients with pertussis and pertussis complications within 90 days of pertussis diagnosis (as shown in the Supplementary, Table S1). We then estimated the number of general practice (GP) visits and associated cost of diagnosed pertussis. We determined cost associated with pertussis and pertussis complication in relation to GP visits, treatment, and additional medication costs among the at-risk population, and estimated the costs using information provided under the Australian government and the Pharmaceutical Benefits Scheme.^18-20^

The costs of general practitioner consultations and pathology services for pertussis cases were estimated, incorporating both out-of-pocket expenses for patients and government contributions. Cost estimates were stratified by healthcare billing status (bulk-billed versus non-bulk-billed), and unit costs sourced from the Australian Government’s Medical Cost Finder.^19,20^ Costs were calculated separately for patients with and without complications, and average annual costs were estimated during the study period.

Analyses were conducted using Microsoft Excel 2010. P-value was calculated using 2-proportion z-test where relevant, and odds ratio was calculated using a two-by-two contingency table and chi-square test. P-value of <0.05 was applied as the statistical significance. This study was approved by the Institutional Review Board (the University of New South Wales (UNSW HREC, Ref. HC220297).

## Results

A total of 764,864 active adult patients were selected for study from 2008 to 2019. Of these, 0.2% (1,788/764,864) were diagnosed with pertussis, 4.1% (31,110/764,864) had undiagnosed coughing illness, and 95% (726,950/764,864) had neither pertussis nor coughing illness (controls) (Figure1).

**Figure 1.**
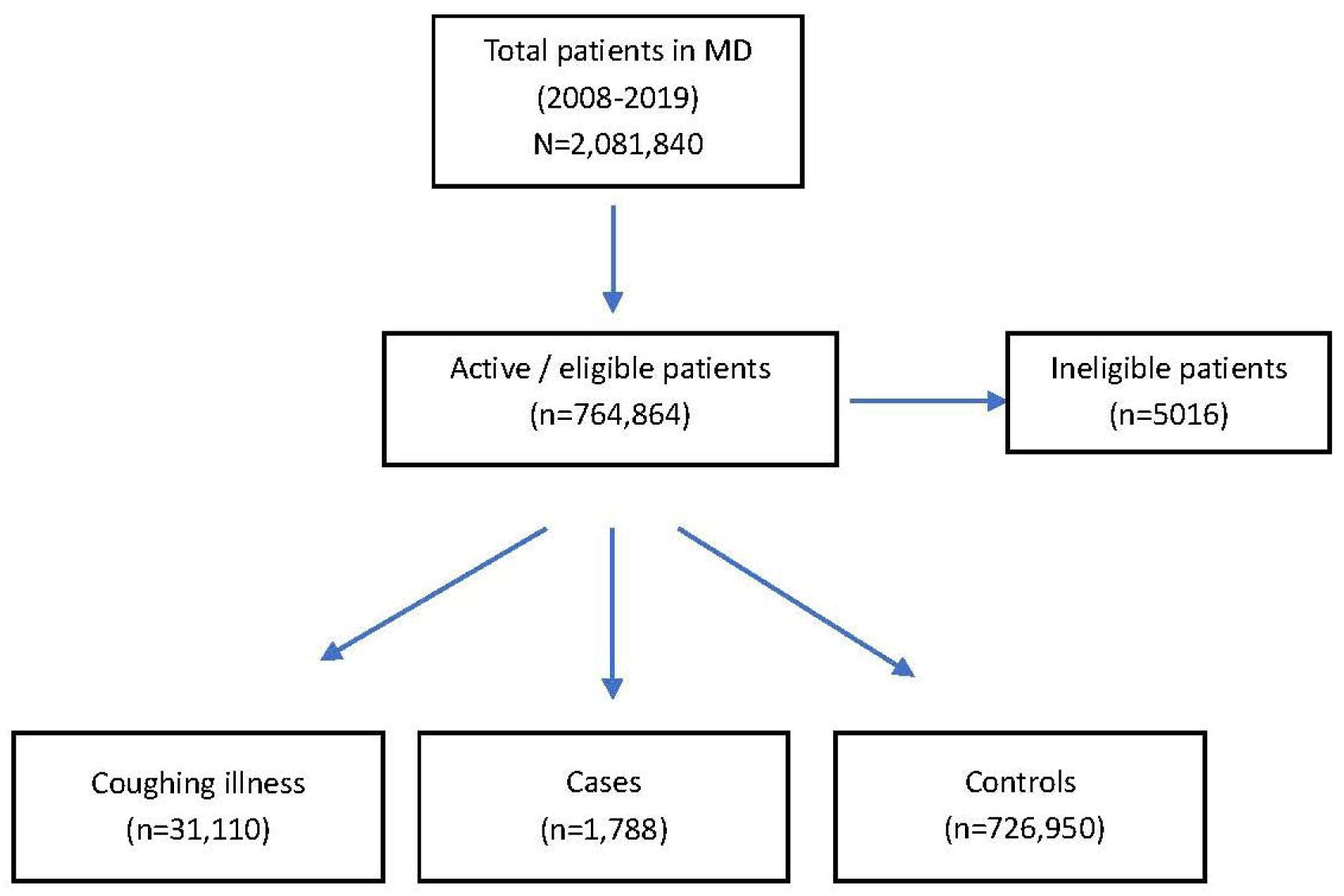
Consort diagram for the study.

Of pertussis cases, 33.7% (602/1,788) were smokers; 58.7% (1,049/1,788) had comorbidities, and 4.4% (78/1,788) of cases had pertussis complications. The overall demographic characteristics of participants are described in Table 1. All measured comorbidities (asthma or COPD, CVD, diabetes and obesity) were significantly higher in pertussis cases compared to controls, with asthma or COPD being the most prevalent comorbidity, occurring in a quarter of pertussis cases.

**Table 1.**
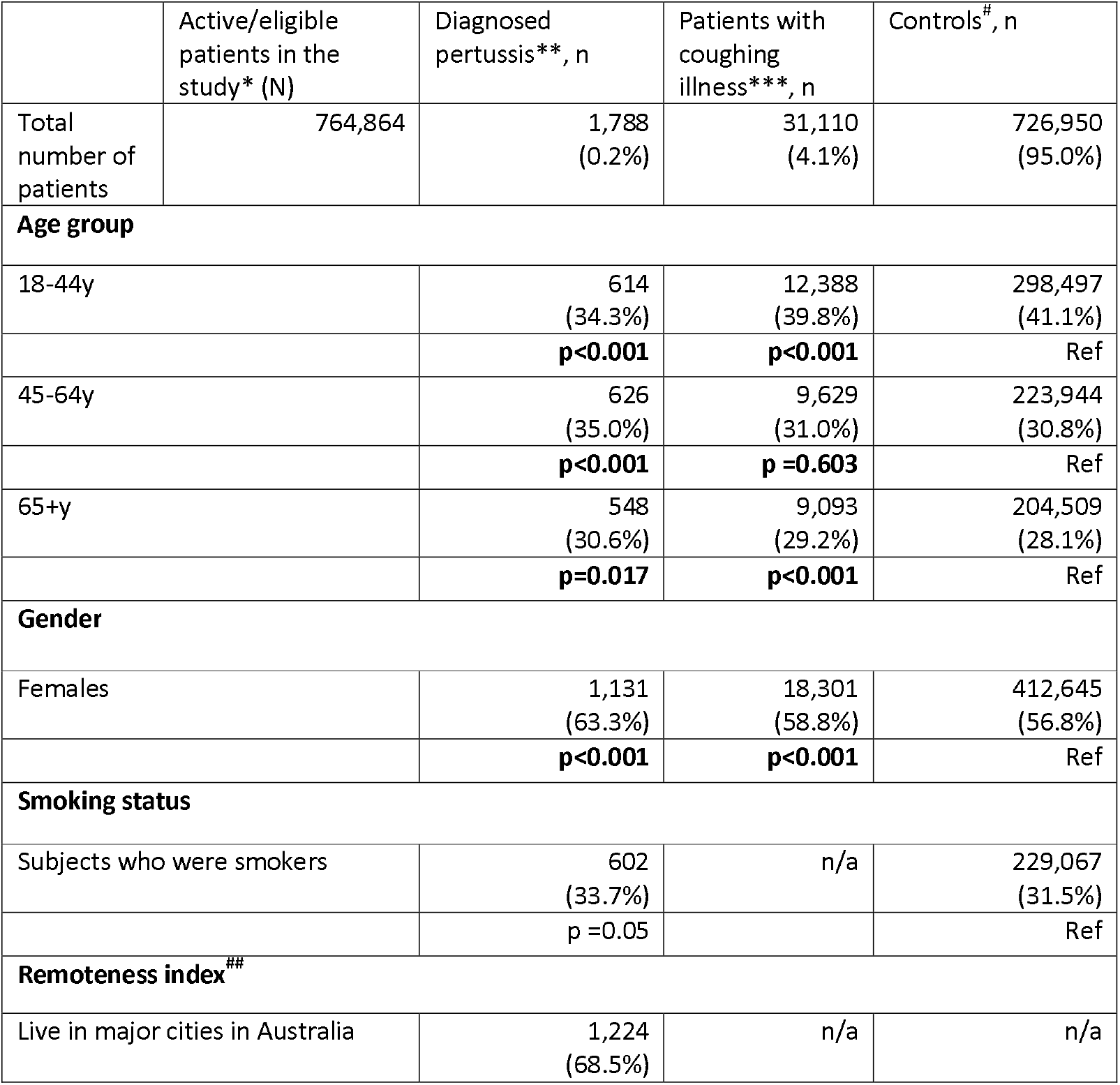
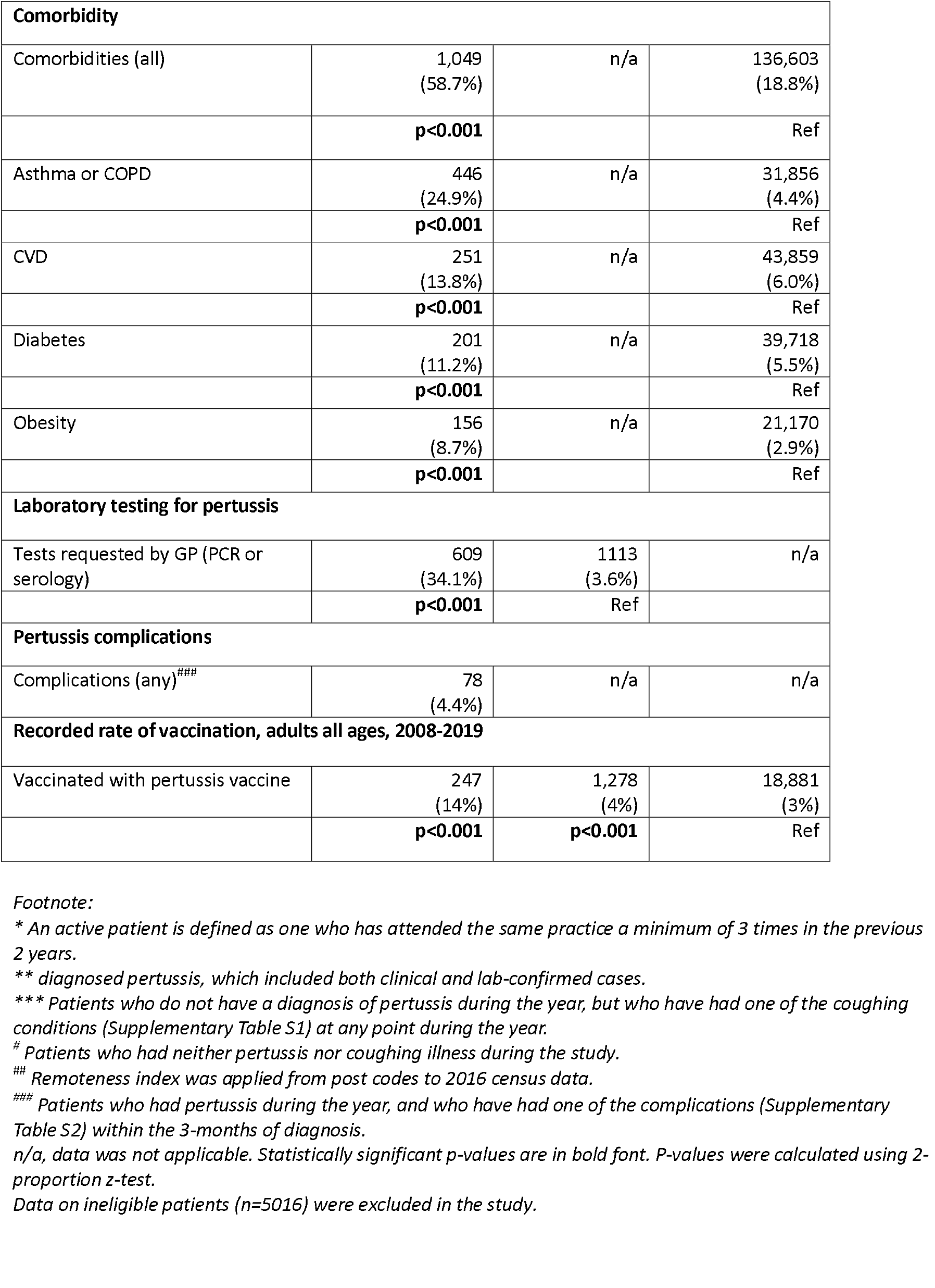
Characteristics of the study population, 2008-2019.

The diagnosis rate of pertussis and coughing illness varied by year but followed a similar trend until the last 3 years, when unspecified coughing illness rose (Figure 2). The all-age average annual diagnosis rate of pertussis was 76.9 per 100,000 population in adults >18 years. The highest rate was recorded in 2011 at 125.1 per 100,000, followed by 102.2 and 94.7 in 2010 and 2015, respectively.

**Figure 2.**
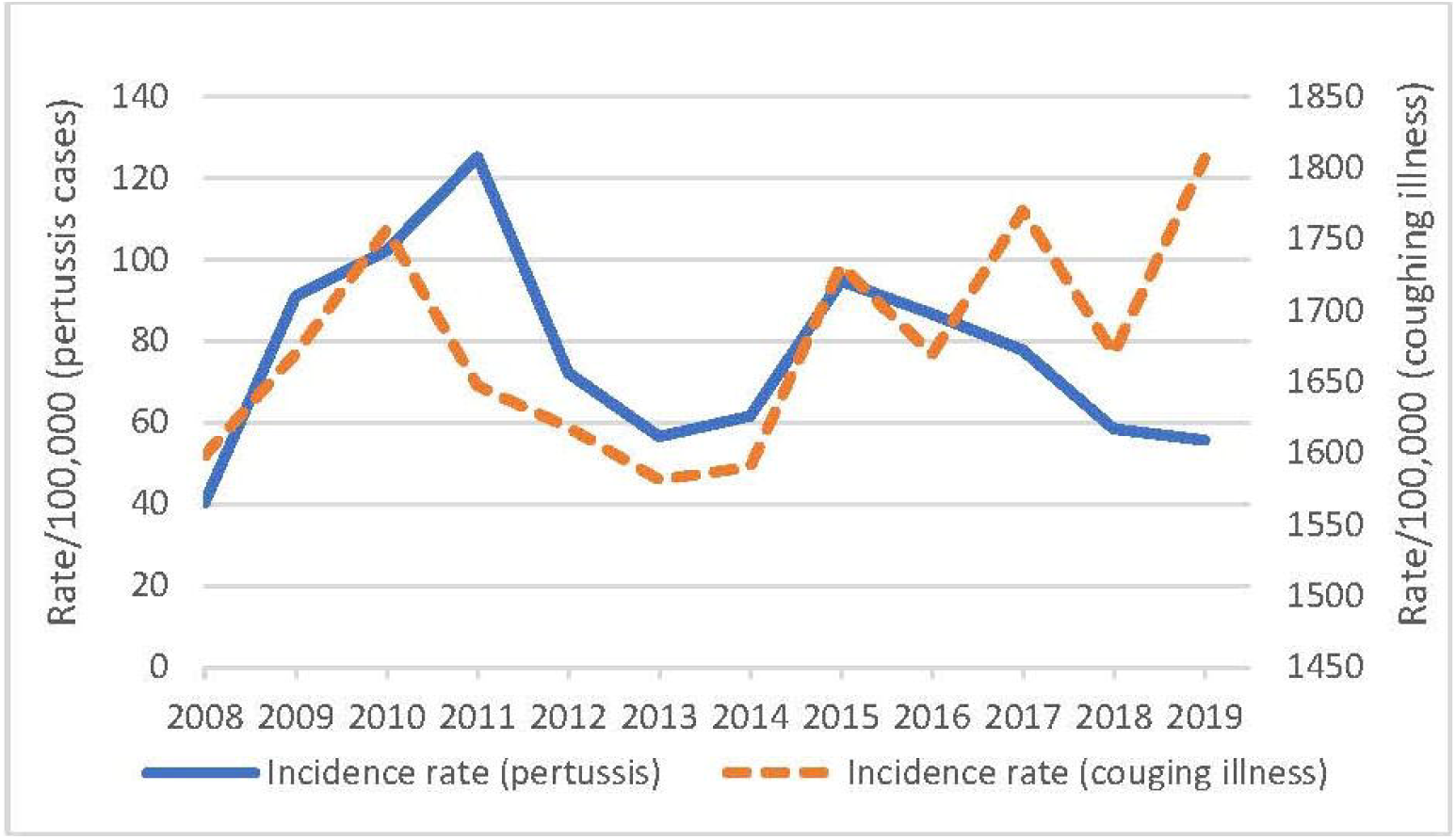
Diagnosis of pertussis and coughing illness rate per 100,000 population, 2008-2019.

The rate of diagnosed pertussis varied by age group as shown in Figure 3, the highest rate was recorded in the 18-44-year age group, followed by the 45–64-year age group in both 2011 and 2015 in the study.

**Figure 3.**
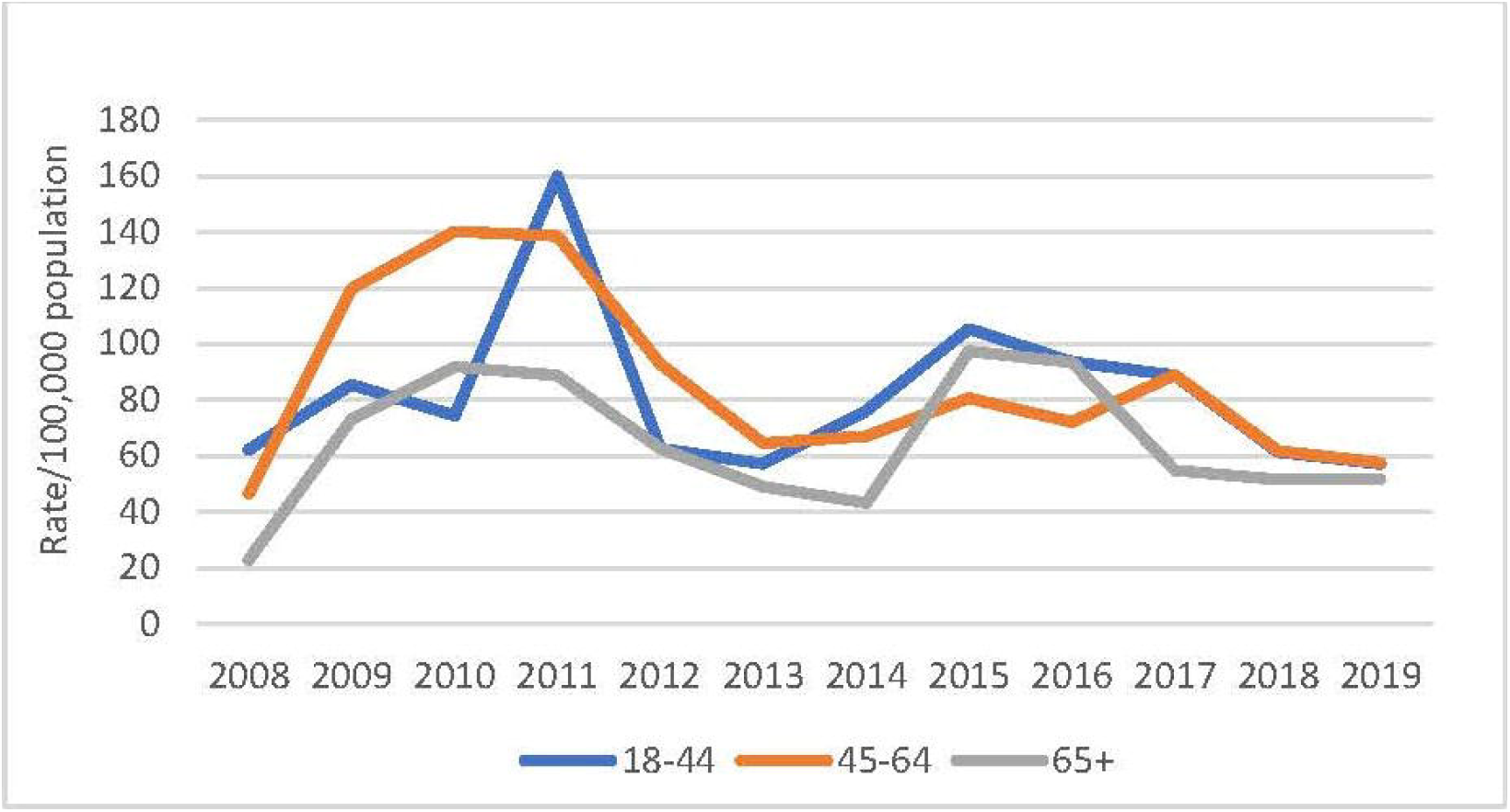
Diagnosis rate of pertussis by age group, 2008-2019.

Among pertussis cases, females had a higher incidence, 63.3% (1,131/1,788) compared to males (36.7%), with an odds ratio (OR) of 1.31 (95%CI 1.19-1.44, p <0.001). There was variation in pertussis diagnosis rates across states and territories. During the study period from 2008-2019, Western Australia had the highest diagnosis rate of pertussis (936.7 per 100,000), followed by New South Wales and Queensland at a rate of 270.3 and 243 per 100,000 population, respectively.

### Chronic conditions or other comorbidities

About 59% of pertussis cases had comorbid conditions such as asthma /COPD or CVD or diabetes or obesity; with asthma/COPD being the most frequent (24.9%), followed by CVD, diabetes and obesity (13.8%, 11.2% and 8.7% respectively). Only 18.8% of controls had comorbidities. Presence of comorbidities among pertussis cases and controls by age group are presented in Table 2.

**Table 2.** Pertussis cases and controls who had comorbidities, by age group, 2008-2019.

| Age Group | # Patients with a diagnosis of pertussis, n | Asthma or COPD, n (%) | p-value | CVD, n (%) | p-value | Diabetes, n (%) | p-value | Obesity , n (%) | p-value |
| --- | --- | --- | --- | --- | --- | --- | --- | --- | --- |
| 18-44y |  |  |  |  |  |  |  |  |  |
| Cases | 614 | 120 (19.5) | <0.001 | * | n/a | 17 (2.8) | 0.014 | 42 (6.8) | <0.001 |
| Controls | 298,497 | 7,466 (2.5) |  | 2,083 (0.7) |  | 4,595 (1.5) |  | 8,440 (2.8) |  |
| 45-64y |  |  |  |  |  |  |  |  |  |
| Cases | 626 | 141 (22.5) | <0.001 | 51 (8.1) | <0.001 | 60 (9.6) | <0.001 | 67 (10.7) | <0.001 |
| Controls | 223,944 | 8,587 (3.8) |  | 8,603 (3.8) |  | 12,788 (5.7) |  | 8,702 (3.9) |  |
| >65y |  |  |  |  |  |  |  |  |  |
| Cases | 548 | 185<br>(33.8) | **<0.001** | 195<br>(35.6) | **<0.001** | 124<br>(22.6) | **<0.001** | 47<br>(8.6) | **<0.001** |
| Controls | 204,509 | 15,803<br>(7.7) |  | 33,173<br>(16.2) |  | 22,335<br>(10.9) |  | 4,028<br>(2.0) |  |

\* values <10 per cell.
Statistically significant p-values are in bold font.

Cases were six times more likely to have had comorbidities compared to controls, with an OR of 6.16 (95%CI: 5.61-6.77), p <0.001. The odds ratio varied with various comorbid conditions among cases and controls, are presented in Supplementary Table S3.

### Pertussis complications

Of cases, 4.4% (78/1,788) had pertussis complications, the most common being pneumonia, ranging from 15-45% of all complications across age groups. The presence of complications across various age groups is shown in Table 3.

**Table 3.** Rate of pertussis complications (any), by age group.

| Age Group | Patients with pertussis, n | Patients with pertussis who had complication/s (any), n | Rate per 1,000 cases | OR 95%CI | p-value |
| --- | --- | --- | --- | --- | --- |
| 18-44y | 614 | 19 | 30.9 | - | Ref |
| 45-64y | 626 | 34 | 54.3 | 1.80 (1.01- 3.19) | **p =0.042** |
| 65+y | 548 | 25 | 45.6 | 1.50 (0.81- 2.75) | p =0.191 |

Statistically significant p-value is in bold font.

The age group 45-64 years had a significantly higher rate of complications, OR=1.80 (95%CI: 1.01-3.19) compared to the 18-44 years. Similarly, the age group >65 years had higher odds of having complications compared to younger cases (18-44 years), with an OR of 1.50 (95%CI: 0.81-2.75), but this was not statistically significant (Table 3).

### Vaccination rate

Of all active patients, 31.3% (239,385/764,864) received any adult vaccination with either influenza, pertussis, pneumococcal or herpes zoster vaccine during the study period.

Of pertussis cases, 13.9% (247 /1,782) were vaccinated against pertussis. Over 90% of cases in the study received their pertussis vaccine within the five-year period (2015-2019). Among the 247 vaccinated cases, 59% (145/247) received the vaccine prior to their pertussis diagnosis. While data were not reported due to data values of <10 per cell, less than 7% of cases received pertussis vaccination after the diagnosis. Notably, pertussis vaccination rates were low in patients with coughing illness and among controls, at 4.1% (1,278/31,254) and 2.6% (18,881/730,094) respectively. Vaccination rates among cases, those with coughing illness and controls by type of vaccine are shown in Figure 4.

**Figure 4.**
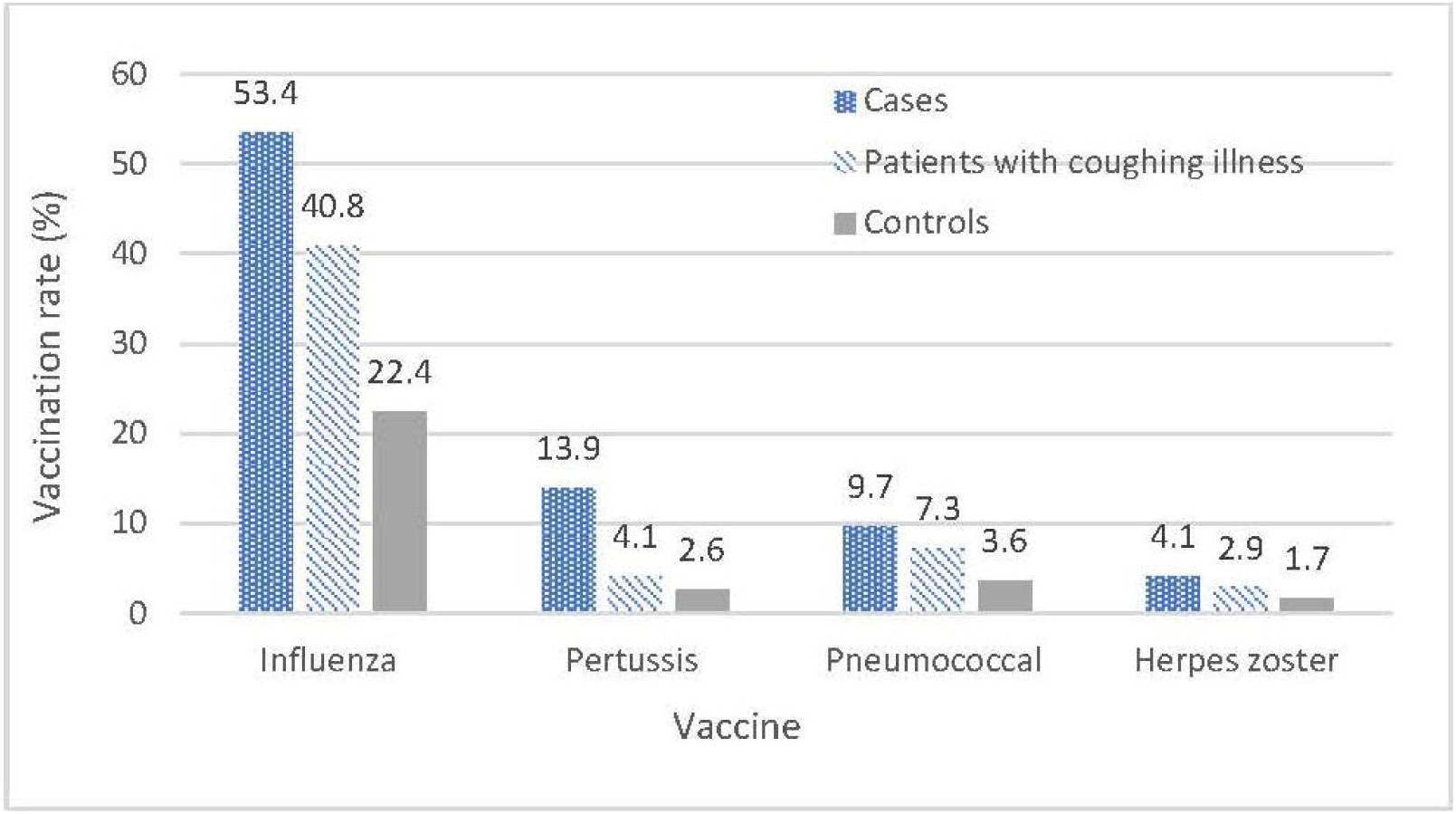
Vaccination rate by group and type of vaccine, 2008-2019, all ages.

Figure 5 shows the rate of vaccination in cases with various vaccines by age group. There was variation in vaccination rates across age groups, and the vaccine uptake was high in older adults >65 years for all vaccines.

**Figure 5.**
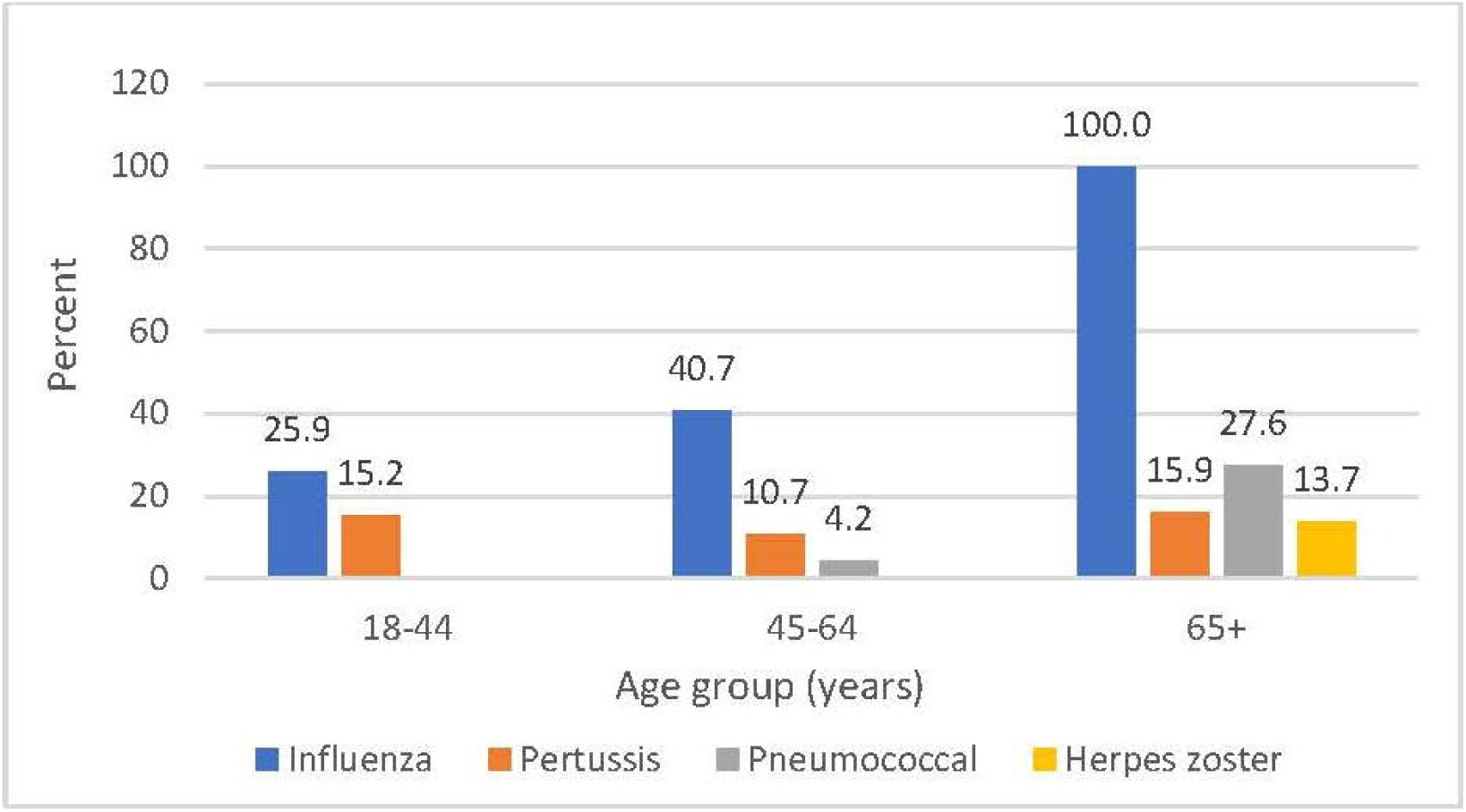
Vaccination rate in cases by age group and type of vaccine, 2008-2019.

### Diagnostic testing

General practitioners requested a total of 831 laboratory tests for XX pertussis cases. There was year-to-year variation in the number of diagnostic tests performed during the study period (Supplementary Figure S1), and the testing trends closely mirrored the number of cases diagnosed over the years. The overall rate of laboratory testing for cases was 34.1%. Among pertussis cases, PCR and serology testing accounted for 57.5% and 42.5% of all tests, respectively. The rate of PCR and serology testing for pertussis among the coughing illness group was 65% (723/1113) and 35% (390/1113) of all tests, respectively. The use of diagnostic methods varied over time; a gradual increase in PCR testing was also observed (Figure 6).

**Figure 6.**
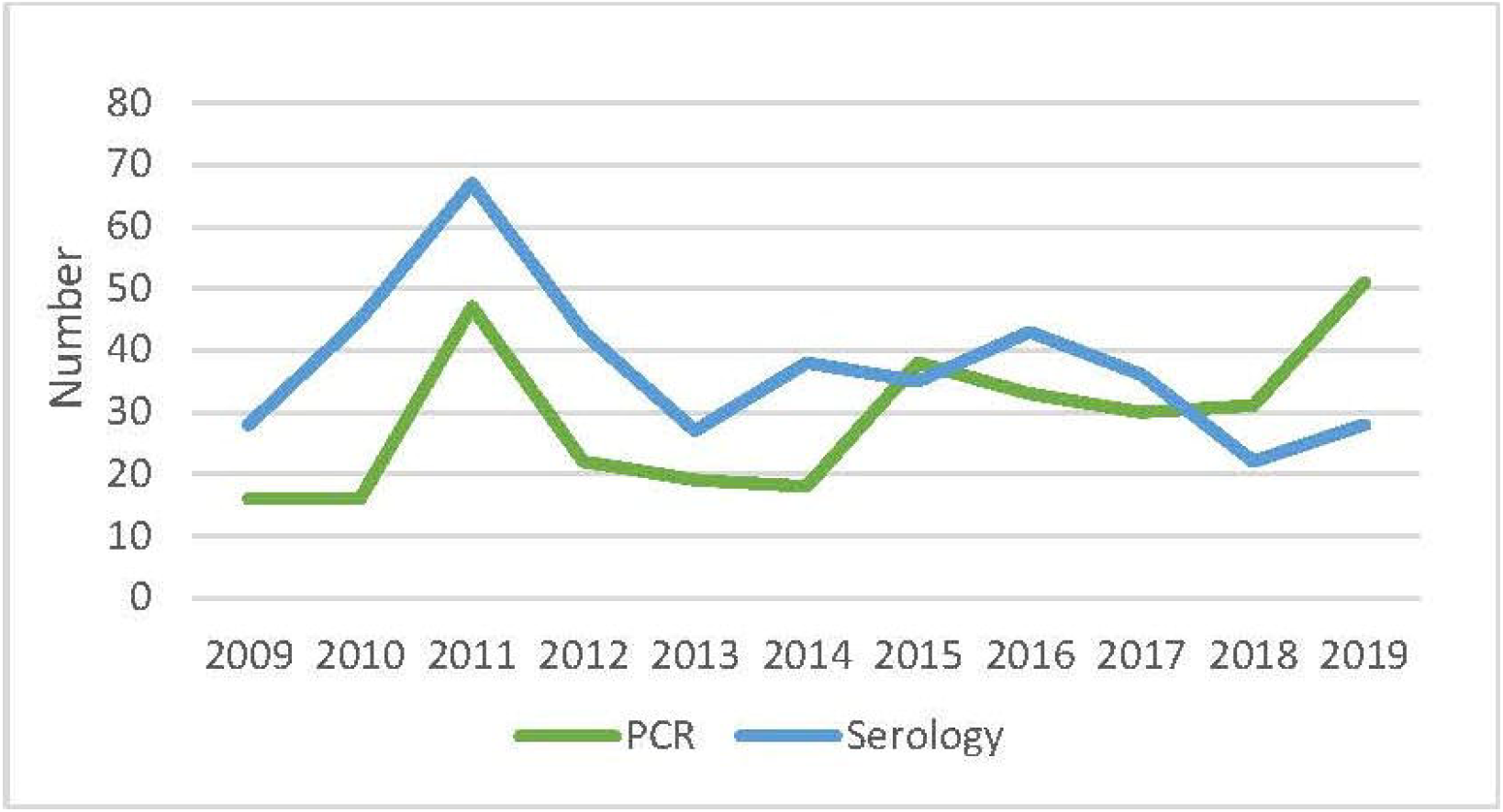
Use of diagnostic method in pertussis cases, 2009-2019. *Footnote:* Year 2008 was omitted due to values of <10 per cell.

### Medications

Table 4 shows the prescription rate per 1,000 patients who had comorbidities. Cases were more likely to have increased use of medications compared to controls who had similar comorbid condition, and that increases with age (data not shown).

**Table 4.** Prescription rate per 1,000 patients, 2008-2019, all ages.

| All ages (2008-2019) |  | Prescription rate per 1,000 among cases | Prescription rate per 1,000 among controls | Rate ratio (Pertussis cases vs controls) |
| --- | --- | --- | --- | --- |
|  | Asthma and COPD medication | 9,269.1 | 6,951.1 | 1.33 (1.29, 1.37) |
|  | CVD medication* | 32,300.8 | 22,646.4 | 1.43 (1.40, 1.46) |
|  | Diabetes medication | 13,189.1 | 7,895.3 | 1.67 (1.61, 1.73) |
|  | Obesity medication | 1,256.4 | 1,157.3 | 1.09 (0.95, 1.25) |

CVD, cardiovascular disease.
\*CVD medication excluded in 18-44y age group, due to values of <10 per cell.

Pertussis cases without complications had an average of nine GP visits related to pertussis, compared to twelve for those with complications. Table 5 presents the associated primary care costs for pertussis cases, including both patient out-of-pocket expenses and government-funded costs. Among patients without complications, the average estimated cost per patient ranged from AUD $473 to $909. For those who developed complications, costs were higher, ranging from AUD $596 to $1,164 per case (Supplementary Table S4). The total estimated average annual cost of pertussis cases analysed in the study was approximately AUD $91,000, with higher overall costs observed among patients who experienced complications.

**Table 5.**
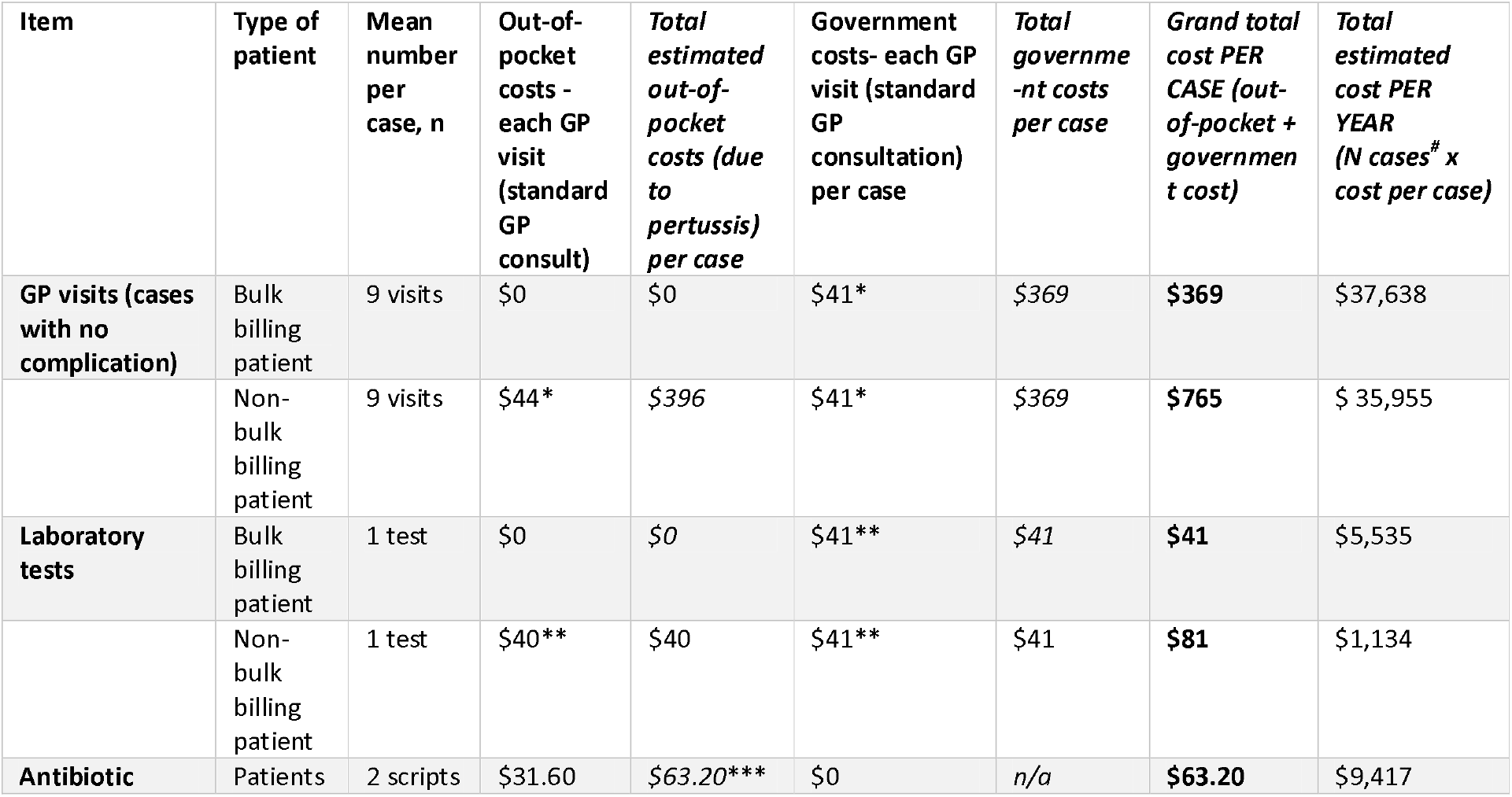
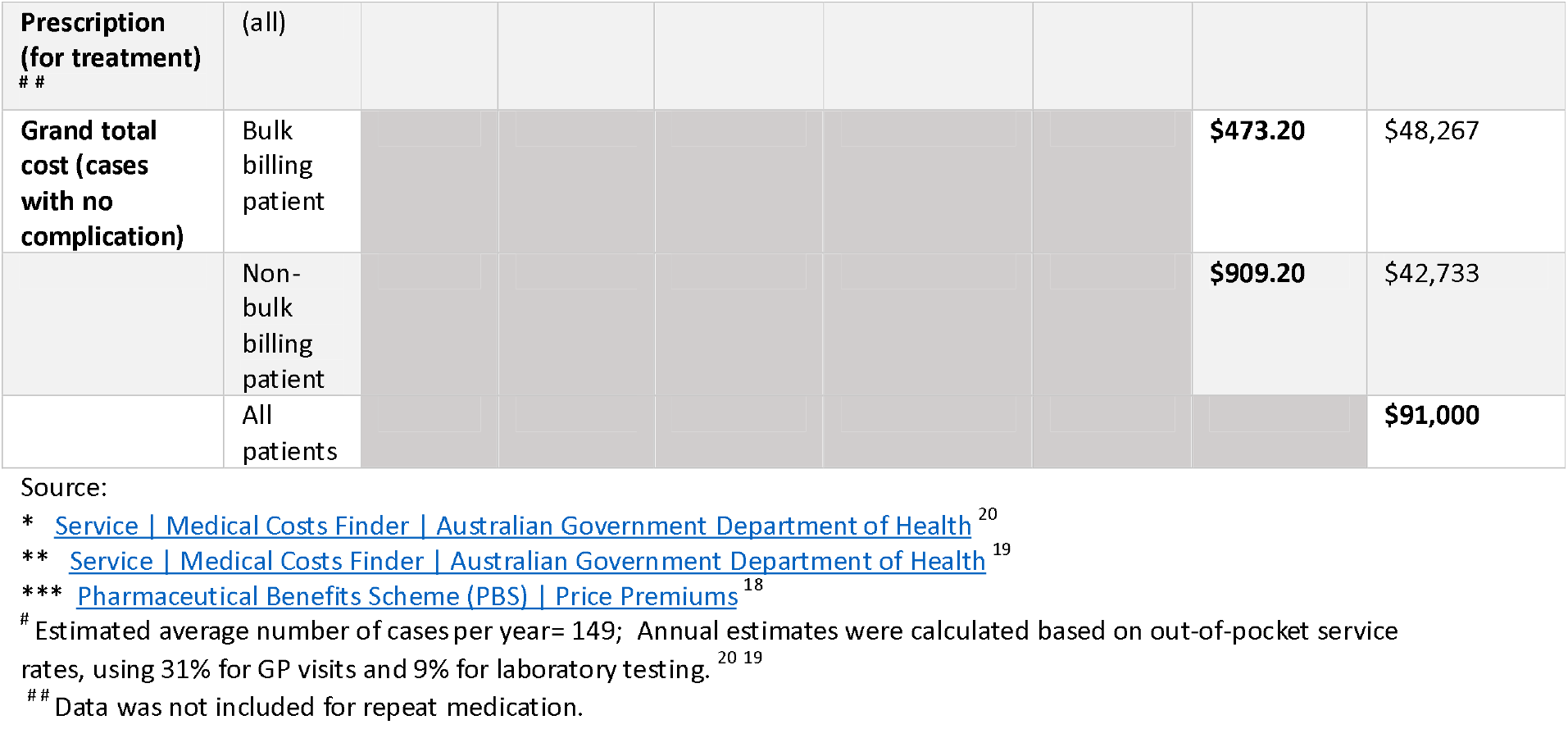
The estimated costs associated with pertussis in adults.

## Discussion

We observed a substantial burden of adult pertussis in the Australian primary care setting over more than a decade, with incidence rates and trends closely aligning with national notification data. The all-age average annual diagnosis rate was 76.9 per 100,000 population, with notable epidemic peaks occurring in 2011 and 2015.^3^ According to the National Notifiable Diseases Surveillance System, the average all-age notification rate of pertussis in Australia was 89.4 per 100,000 population between 2008 and 2019.^5^ Similarly, an observational study conducted in Australian primary care from 2015 to 2019 reported pertussis incidence rates ranging from 57.6 to 91.4 per 100,000 among adults aged 50 years and older.^21^ A descriptive study from the national surveillance program in Australia found that the average rate of pertussis in adults aged 65 years and over was 86 per 100,000 population from 2006-2012.^22^ Another large population-based cohort study from the state of New South Wales reported a pertussis incidence rate of 94 per100,000 person-years in adults aged ≥45 years.^23^ Like several other studies, we found that females had a higher incidence of pertussis.^3,21,23^ The higher rate in women may reflect greater contact with children due to parental and grandparental care.^24,25^ Like Australia, studies from other countries showed an increasing burden of pertussis in adults in recent years.^1,26,27^ There was some variation in diagnosis rate of pertussis by states and territories, with Western Australia having the highest rate during the study period, which may reflect the increasing burden of disease or be due to increase in testing practices in that state.

Asthma and COPD are one of the most common chronic conditions in Australia, and people with asthma /COPD are at increased risk of pertussis infection and its complications.^1,2,28,29^ One Australian study found that the rate of reported pertussis infection in asthmatic adults aged ≥50 years was 160 per 100,000 compared to 90 per 100,000 person-years in those who did not have asthma.^23^ A history of tobacco smoking and asthma was likely to increase the risk of pertussis hospitalisation in adults aged >65 years.^30^ Asthma, COPD, immunosuppression, obesity and smoking are shown in other studies to be risk factors for pertussis.^31^

Severe pertussis in adults can result in complications such as pneumonia, sinusitis, seizures, urinary incontinence, encephalitis, secondary respiratory infection, rib fracture, subdural haemorrhage and even death.^11,27,32^ We showed that less than five percent of pertussis cases in primary care developed complications, most commonly pneumonia. However, we did not capture data on hospitalization or emergency department visits, which may be the first site of presentation for people with severe complications. This may explain why people aged >65 years had a lower rate of complications compared to 45-64 years-old in our study. This may be due to fewer presentations to GPs among older cases, who may have presented to emergency departments instead.^30^

Over 85% of patients with pertussis were unvaccinated, despite seeing a GP at least three times over two years, highlighting a critical missed opportunity for prevention. We observed low pertussis vaccination rates, even among older patients and those with comorbidities. According to the Australian Immunisation Register, only about 21 % of Australian adults aged 50 years and older were vaccinated against pertussis in 2023.^13^ Tetanus boosting is an important opportunity for pertussis vaccination. A recent study found that tetanus vaccination rate of 30%, compared to just 20% for pertussis, which likely reflects use of dT instead of TdaP for tetanus boosters.^13^ In a 2009 vaccination survey, pertussis vaccine coverage in Australian adults was only 11.3%.^33^ In this primary care setting, we found that only 11% of cases aged 40-64 years and 16% of those 65 years and over had received a pertussis vaccine. The majority of vaccinated cases had received the vaccine within the past 5-years (2015-2019); notably, where 59% of cases were vaccinated prior to their pertussis diagnosis, while approximately 35% received the vaccine or were recommended vaccination only after their diagnosis. Low adult pertussis vaccine uptakes have been reported in other Australian studies.^12,21^ The slightly higher vaccination rates observed among pertussis cases compared to controls may reflect a healthy user bias, where patients with chronic conditions or those who are at higher risk of pertussis are more likely to seek health care services and receive vaccinations. However further investigation is warranted to better understand this association.

Vaccine efficacy for acellular pertussis vaccine in adults 15-65 years in a randomized, controlled trial in US found a protection rate of 92%,^34^ and a meta-analysis of pertussis vaccine in adolescents and adults mentioned a short-term effectiveness of acellular pertussis vaccine with vaccine response rates of >85% among the studies.^35^ However, a low to moderate vaccine effectiveness (VE) of pertussis was reported in adults in studies.^36,37^ Vaccine effectiveness can be dependent on age and study setting and time since vaccination. A moderate vaccine effectiveness, an adjusted VE of 52% was also noted in an Australian study for PCR-confirmed cases.^36^ Authors of that study also reported a variation in VE by timing of vaccination, and a non-significant VE of 42% (-23 to 73%) was found in cases if vaccinated between 2-5 years. More research is needed to evaluate the effectiveness of pertussis vaccine in adults. Recommendations for adult pertussis vaccination vary across countries.^31^ Nonetheless, in settings with a high disease burden, even a vaccine with moderate effectiveness can have a substantial impact on population health.^38^

In addition to health impacts, pertussis results in increased use of health services and an economic burden. In a UK study, in adults aged ≥50 years, individuals with asthma had a higher health care utilisation and direct medical care costs in individual was increased by £370 (95% CI 128–650; 26.4% higher) compared to people without asthma in the study.^1^ Our costing did not include hospital costs, which are far higher than community costs per episode. Nonetheless, during an epidemic year, community costs of pertussis may be substantial. In 2024, Australia recorded the largest epidemic of pertussis with over fifty-seven thousand notified cases in the year.^5^ If all these cases had seen a GP, based on our estimated costs per episode, the total cost incurred in primary care may have been at least around $35 million dollars, excluding any hospitalisation costs.

The laboratory methods used to diagnose pertussis has varied over time. Consistent with our findings, the use of PCR has ranged from 50% to 90% of all tests compared to serology, which varied from 50% to 10% between 2014 and 2018 in Australia.^3^ Availability and use of PCR testing for pertussis have been increased significantly since 2005, following the introduction of public funding for laboratories conducting PCR tests under the Medicare Benefit Schedule.^39^ However, diagnostic testing for pertussis also depends on GP’s recognition or awareness of pertussis as a differential diagnosis. Due to under-ascertainment and underreporting, the true burden of pertussis in adults may be higher. Pertussis can present with chronic or prolonged cough and is frequently under-ascertained or misdiagnosed in adult patients.^40^ We found that 4.1% of patients had an unspecified coughing illness during the study period and the majority had not been tested for pertussis. Improved awareness and testing for pertussis is needed. Another area for research is the burden of pertussis in aged care settings, where pertussis epidemics do occur, however testing in these settings is mostly limited to viral PCR, and awareness of pertussis may be even lower than in the community.^41^

There were several limitations in the study. First, due to privacy, data were not provided in cells with values less than 10, resulting in missing data. We could not obtain specific data on complications, because the occurrence of individual complications other than pneumonia was less than 10 counts. Second, data on pathology test results for laboratory confirmation of cases could not be extracted by MD software, so we had to use tests ordered as a proxy. Third limitation was that when an eligible patient attended two practices, the patient was counted as two distinct patients; thus, this could lead to double counting of active patients in the study population. Fourth, depending on the data we received in the study, multivariable analyses could not be done using advanced analyses to control confounding variables or inherent sociodemographic differences between cases and controls were not possible. Odds ratio presented should interpret with caution. Also, measures of vaccine protection or effectiveness should not be estimated from this study, as data were provided in aggregated form. Fifth, due to limitation of data in our study, cost estimations were restricted to primary care related to pertussis, which reflects a lower cost compared to hospitalisation. Finally, hospitalisations, which represent the most severe end of the spectrum of pertussis, were not captured in the MD data.

## Conclusions

We found a substantial burden of pertussis in adults in primary care, with higher incidence in people with comorbidities, all of which causes a health and economic burden to Australia, especially during high epidemic years. Asthma and COPD are the most important comorbid conditions in people with pertussis. Pertussis vaccine uptake in the primary care setting was very low, reflecting a missed opportunity for prevention. Diagnostic testing for pertussis was also low, and a proportion of undiagnosed coughing illness may be caused by pertussis, especially in epidemic years. GPs play an important role in vaccination, awareness and testing for pertussis.

## Data Availability

Data presented in the study may be available upon reasonable request to the authors.

## Informed Consent Statement

Not applicable. This study collected de-identified, aggregated patient records data in the MedicalDirector from the participating clinics.

## Acknowledgements

The authors thank Angus Cuskelly (BCompSci), Tim Parker (BSc) and Charmaine Tam (PhD) from MedicalDirector for their assistant with data extraction for this study.

