## Supplementary Tables for "The burden of pertussis disease and vaccination coverage in Australian adults attending primary health care"

Table S1. Lists of coughing illnesses

Chronic cough

Bacterial bronchitis

Chronic bronchitis

Eosinophilic bronchitis

Recurrent bronchitis

Complicated bronchitis

Viral bronchitis

Laryngotracheobronchitis

Post bronchitis cough

Chronic Sino-bronchitis

Post infective cough

Post bronchitis cough

Croupy cough

Post viral cough

Wheezy bronchitis

Table S2. Lists of complications

Encephalitis

Seizures

Pneumonia

Rib fracture

Subdural haemorrhage

Syncope

Urinary incontinence

Rectal prolapse

Table S3. Number of patients with comorbidities and rate per 1,000 among cases and controls, all ages.

|  | Asthma or COPD | | CVD | | Diabetes | | Obesity | |
| --- | --- | --- | --- | --- | --- | --- | --- | --- |
|  | n, (Rate /1,000  Patients) | OR  95%CI, p-value | n,  (Rate /1,000  Patient) | OR  95%CI, p-value | n,  (Rate /1,000  Patients) | OR  95%CI, p-value | n,  (Rate /1,000  Patients) | OR  95%CI, p-value |
| Pertussis cases | 446 (249.4) | ***-*** | 246 (140.4) | ***-*** | 201  (112.4) | ***-*** | 156  (87.3) | ***-*** |
| Controls | 31,856 (43.8) | ***-*** | 43,859  (60.3) | ***-*** | 39,718  (54.6) | ***-*** | 21,170  (29.1) | ***-*** |
| Cases vs controls |  | OR=7.25 (6.51- 8.08)  **<0.001** |  | OR=2.54 (2.22- 2.91)  **<0.001** |  | OR=2.19 (1.89- 2.54)  **<0.001** |  | OR=3.19 (2.70- 3.76)  **<0.001** |

*Footnote:*

OR, odds ratio

Statistically significant p-values are in bold font.

Table S4. The estimated costs associated with pertussis in adults who had complications

| Item | Type of patient | Mean number per case, n | Out-of-pocket costs - each GP visit (standard GP consult) | *Total estimated out-of-pocket costs (due to pertussis) per case* | Government costs- each GP visit (standard GP consultation) per case | *Total government costs per case* | *Grand total cost PER CASE (out-of-pocket + government cost)* |
| --- | --- | --- | --- | --- | --- | --- | --- |
| GP visits (cases with complications) | Bulk billing patient |  | $0 | $0 | $41 | *$492* | *$492* |
|  | Non-bulk billing patient | 12 visits | $44 | *$528* | $41 | *$492* | $1,020 |
| Laboratory tests | Bulk billing patient |  | $0 | *$0* | $41** | *$41* | $41 |
|  | Non-bulk billing patient | 1 test | $40 | $40 | $41 | $41 | $81 |
| Antibiotic Prescription (for treatment) ^#^ ^#^ | Patients (all) | 2 scripts | $31.60 | *$63.20**** | $0 | *n/a* | $63.20 |
| Grand total cost (cases with complications) | Bulk billing patient |  |  |  |  |  | **$596.20** |
|  | Non-bulk billing patient |  |  |  |  |  | **$1,164.20** |

Source:

*	[Service | Medical Costs Finder | Australian Government Department of Health](https://medicalcostsfinder.health.gov.au/service/?id=Q23&mode=OH&specialtyname=General%20practice%20(GP)&specialty=019999&tab=fees) ^20^

**	[Service | Medical Costs Finder | Australian Government Department of Health](https://medicalcostsfinder.health.gov.au/service/?id=Q23&mode=OH&tab=fees&specialty=021701&specialtyname=Pathology) ^19^

*** [Pharmaceutical Benefits Scheme (PBS) | Price Premiums](https://www.pbs.gov.au/browse/brand-premium) ^18^

^#^ Estimated average number of cases per year= 149; ^# #^ Data was not included for repeat medication
