## Supplementary figures and images for "The burden of pertussis disease and vaccination coverage in Australian adults attending primary health care"

### Supplementary figure

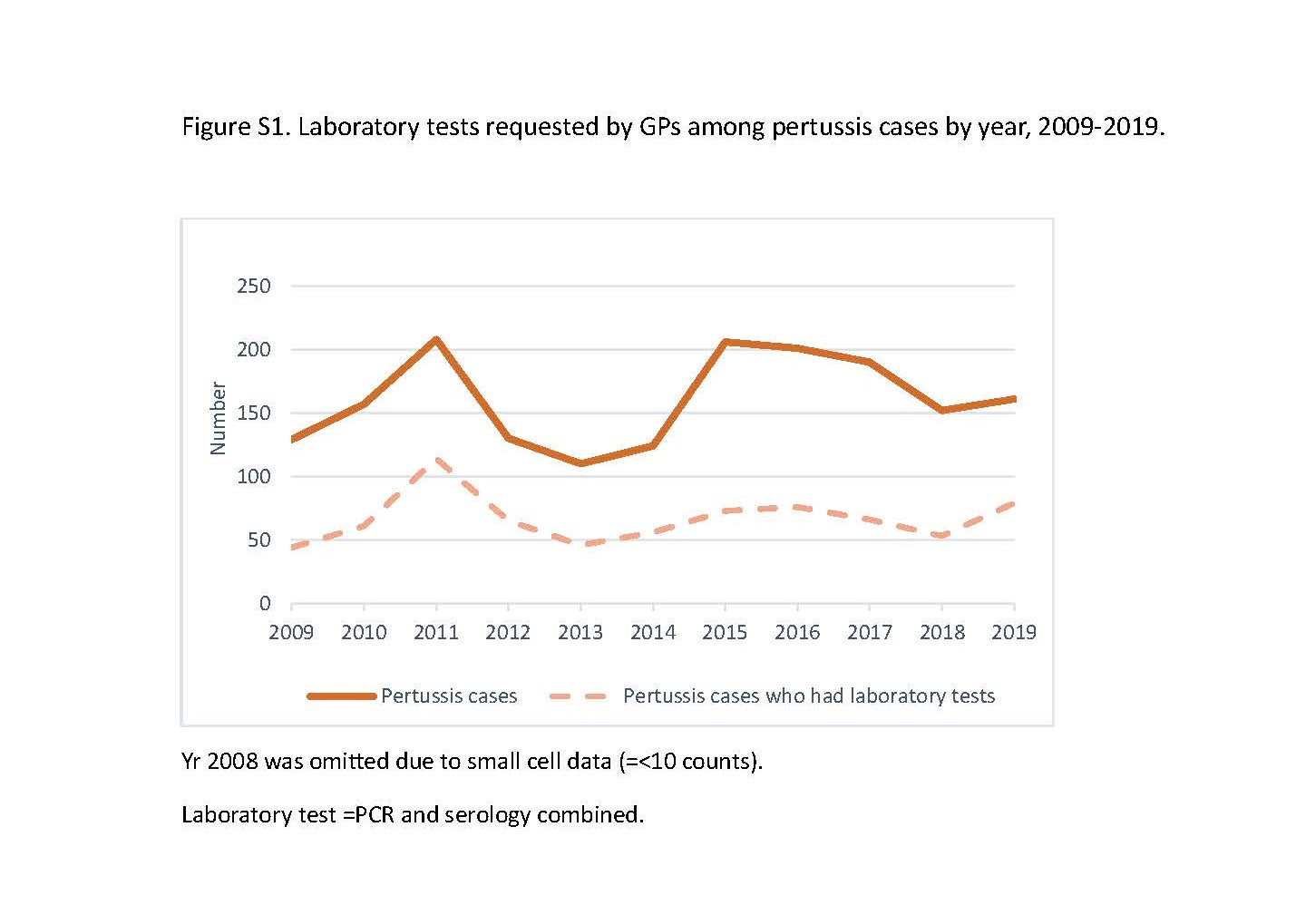
